# Intra-pulmonary and intra-cardiac shunts in adult COVID-19 versus non-COVID ARDS ICU patients using echocardiography and contrast bubble studies (COVID-Shunt Study): a prospective, observational cohort study

**DOI:** 10.1101/2022.08.04.22278445

**Authors:** Vincent I. Lau, Graham D. Mah, Xiaoming Wang, Leon Byker, Andrea Robinson, Lazar Milovanovic, Aws Alherbish, Jeffrey Odenbach, Cristian Vadeanu, David Lu, Leo Smyth, Mitchell Rohatensky, Brian Whiteside, Phillip Gregoirev, Warren Luksun, Sean van Diepen, Dustin Anderson, Sanam Verma, Jocelyn Slemkov, Peter Brindley, Demetrios J. Kustogiannis, Michael Jacka, Andrew Shaw, Matt Wheatley, Jonathan Windram, Dawn Opgenorth, Nadia Baig, Oleksa G. Rewa, Sean M. Bagshaw, Brian M. Buchanan

**Affiliations:** Department of Critical Care Medicine, Faculty of Medicine and Dentistry, University of Alberta, and Alberta Health Services, Edmonton, Alberta, Canada; Health Services Statistical and Analytic Methods, Alberta Health Services, Edmonton, Alberta, Canada; Division of Cardiology, Department of Medicine, Faculty of Medicine, and Alberta Health Services, Edmonton, Alberta, Canada; Department of Critical Care Medicine, Cumming School of Medicine, University of Calgary, Calgary, Alberta, Canada; Department of Emergency Medicine, Faculty of Medicine and Dentistry, University of Alberta, Edmonton, Alberta, Canada; Faculty of Medicine and Dentistry, University of Alberta, Edmonton, Alberta, Canada; Department of Medicine, Cumming School of Medicine, University of Calgary, Calgary, Alberta, Canada; Department of Anesthesiology & Pain Medicine, Faculty of Medicine and Dentistry, University of Alberta, Edmonton, Alberta, Canada; Division of Neurology, Department of Medicine, Faculty of Medicine and Dentistry, University of Alberta, Edmonton, Alberta, Canada; Department of Intensive Care and Resuscitation, Cleveland Clinic, Cleveland, Ohio, United States of America; Department of Neurosurgery, Faculty of Medicine and Dentistry, University of Alberta, Edmonton, Alberta, Canada; School of Public Health, University of Alberta, Edmonton, Alberta, Canada

**Keywords:** mortality, COVID-19, pandemic, shunt, hypoxemia, bubble study, transcranial Doppler, echocardiography

## Abstract

**Importance:** Studies have suggested intra-pulmonary shunts may contribute to hypoxemia in COVID-19 ARDS and may be associated with worse outcomes.

**Objective:** To evaluate the presence of right-to-left (R-L) shunts in COVID-19 and non-COVID ARDS patients using a comprehensive hypoxemia work-up for shunt etiology and associations with mortality.

**Design, Setting, Participants:** We conducted a multi-centre (4 Canadian hospitals), prospective, observational cohort study of adult critically ill, mechanically ventilated, ICU patients admitted for ARDS from both COVID-19 or non-COVID (November 16, 2020-September 1, 2021).

**Intervention:** Contrast-enhanced agitated-saline bubble studies with transthoracic echocardiography/transcranial Doppler (TTE/TCD) ± transesophageal echocardiography (TEE) assessed for the presence of R-L shunts.

**Main Outcomes and Measures:** Primary outcomes were shunt incidence and association with hospital mortality. Logistic regression analysis was used to determine association of shunt presence/absence with covariables.

**Results:** The study enrolled 226 patients (182 COVID-19 vs. 42 non-COVID). Median age was 58 years (interquartile range [IQR]: 47-67) and APACHE II scores of 30 (IQR: 21-36). In COVID-19 patients, the incidence of R-L shunt was 31/182 patients (17.0%; intra-pulmonary: 61.3%; intra-cardiac: 38.7%) versus 10/44 (22.7%) non-COVID patients. No evidence of difference was detected between the COVID-19 and non-COVID-19 shunt rates (risk difference [RD]: -5.7%, 95% CI: -18.4-7.0, p=0.38). In the COVID-19 group, hospital mortality was higher for those with R-L shunt compared to those without (54.8% vs 35.8%, RD: 19.0%, 95% CI 0.1-37.9, p=0.05). But this did not persist at 90-day mortality, nor after regression adjustments for age and illness severity.

**Conclusions:** There was no evidence of increased R-L shunt rates in COVID-19 compared to non-COVID controls. Right-to-left shunt was associated with increased in-hospital mortality for COVID-19 patients, but this did not persist at 90-day mortality or after adjusting using logistic regression.

**Key Points:** 

**Question:** Does right-to-left shunt incidence increase with COVID-19 ARDS compared to non-COVID, and is there association with shunt incidence and mortality?

**Findings:** In this prospective, observational cohort study, we showed no statistically significant difference in shunt prevalence between COVID-19 ARDS patients (17.0%) and non-COVID patients (22.7%). However, in COVID-19 patients, there was a difference in hospital mortality for those with shunt (54.8%) compared to those without shunt (35.8%), but this difference did not persist at 90-day mortality, nor after regression adjustments for age and illness severity.

**Meaning:** There was no evidence of increased R-L shunt rates in COVID-19 compared to non-COVID or historical controls. Right-to-left shunt presence was associated with increased hospital mortality for COVID-19 patients, but this did not persist for 90-day mortality or after adjustment using logistic regression.

## Background

Severe acute respiratory syndrome coronavirus 2 (SARS-CoV-2), or coronavirus disease-19 (COVID-19), has infected at least 500 million people, and killed over 6 million. The primary cause of death is usually intractable hypoxemia from acute respiratory distress syndrome (ARDS).^1^ However, some literature raised the possibility of other causes of hypoxemia: specifically right-to-left shunts.^2^ Autopsies from COVID-19 pneumonia patients also demonstrated pulmonary capillary deformations,^3^ and dual-energy computed tomography images (CTs) suggested pulmonary vessel dilatation.^4^

A recent study reported a right-to-left (R-L) shunt in 83% of adult ICU patients with severe COVID-19.^5^ The authors concluded this was secondary to increased pulmonary vascular dilation. However, sample size was small (n=18) and they relied upon agitated-saline microbubbles via transcranial Doppler (TCD) of the bilateral middle cerebral arteries.^6^ However, they could not rule out intra-cardiac disease, as neither transthoracic nor transesophageal echocardiography (TTE/TEE) was performed.^6^ This incidence of shunt was significantly higher than historical ARDS controls,^5,7^ it raised the possibility that COVID-19 ARDS might be associated with increased R-L shunt.

In contrast, another study reported lower rates of shunt in COVID-19 ARDS patients: 10% with PFO, and 20% with detectable transpulmonary bubble transit,^8^ more in-line with historical controls.^7,9^ However, numbers were relatively low (n=60) and the study used contrast-enhanced TTE (but not TEE). While an improvement on TCD, TTE lacks sufficient sensitivity to fully assess the atrial septum.^8,10,11^

A recent systematic review suggested an association between R-L shunts and increased mortality.^12^ Therefore, the purpose of our study was to compare COVID-19 ARDS and non COVID-19 ARDS ICU patients for R-L shunt presence, shunt etiology (intra-pulmonary/intra-cardiac), and associations with mortality. We utilized a comprehensive hypoxemia protocol that included contrast-enhanced TTE/transcranial Doppler (TCD) and TEE.

## Methods

This study was reviewed and fully approved by the local institutional review board (University of Alberta Research Ethics Board: PRO00104364). Waived consent for data was obtained given that TTE, TCD, and TEE are all within standard of care for severe hypoxemia at our institution (and de-identified registry data was available for all patients). Clinical assent/consent for TEE was obtained from either the patient’s substitute-decision maker and/or attending physician.

### Setting and Study Design

Four Canadian intensive care units (ICUs) in Edmonton, Alberta, Canada participated in this prospective, observational cohort study. All are tertiary care referral centres, caring for complex medical, trauma, surgical, oncologic, and transplant patients. All sites are equipped with portable ultrasound machines (Fujifilm Sonosite, Bothell, WA, USA) with probes for TTE, TEE and TCD. Following a POCUS study by the physician, images are saved and automatically uploaded to the Qpath TM (Telexy, Maple Ridge, BC, Canada) archiving system, along with a report charted from the scanning physician.

We recruited eligible consecutive patients between November 16, 2020, to September 1, 2021. Patients were included if they were diagnosed with ARDS who were receiving invasive mechanical ventilation plus COVID-19 pneumonia (comparator group) vs. ARDS without COVID (control group). Patients were excluded if <18 years old.

### Working definitions

We defined COVID-19 infection as having a polymerase-chain reaction (PCR) nucleic acid test (NAT) confirmed by healthcare-approved assay [ref].

We used the 2012 Berlin ARDS definition.^13^ We defined and diagnosed R-L intrapulmonary shunts and intra-cardiac shunts as per the American Society of Echocardiography, where intra-cardiac shunt was defined as a positive bubble study usually within 1-2 cardiac cycles, and evidence of PFO/ASD via TTE or TEE with colour Doppler.^11,14^ An intra-pulmonary shunt was defined as evidence of positive bubble study usually within 4-8 cardiac cycles, with no evidence of PFO/ASD on a TTE or TEE with colour Doppler.^11,14^

A positive TCD study was defined by detection of any microbubbles during insonation of the middle cerebral artery with pulse-wave Doppler and injection of agitated-saline contrast with and without simulated Valsalva (simulating increased intra-abdominal pressure by pressing on the abdomen, and then releasing). We did not categorize severity, only the binary presence/absence of a R-L shunt by TCD.^15^

### Hypoxemia shunt workup

We performed an intra-cardiac and intra-pulmonary shunt workup for hypoxemia in COVID-19 pneumonia patients. All operators and sonographers wore full personal protective equipment. The shunt bubble study protocol is further outlined in Figure 1, including full explanations of TTE/TCD/TEE protocols.

**Figure 1.**
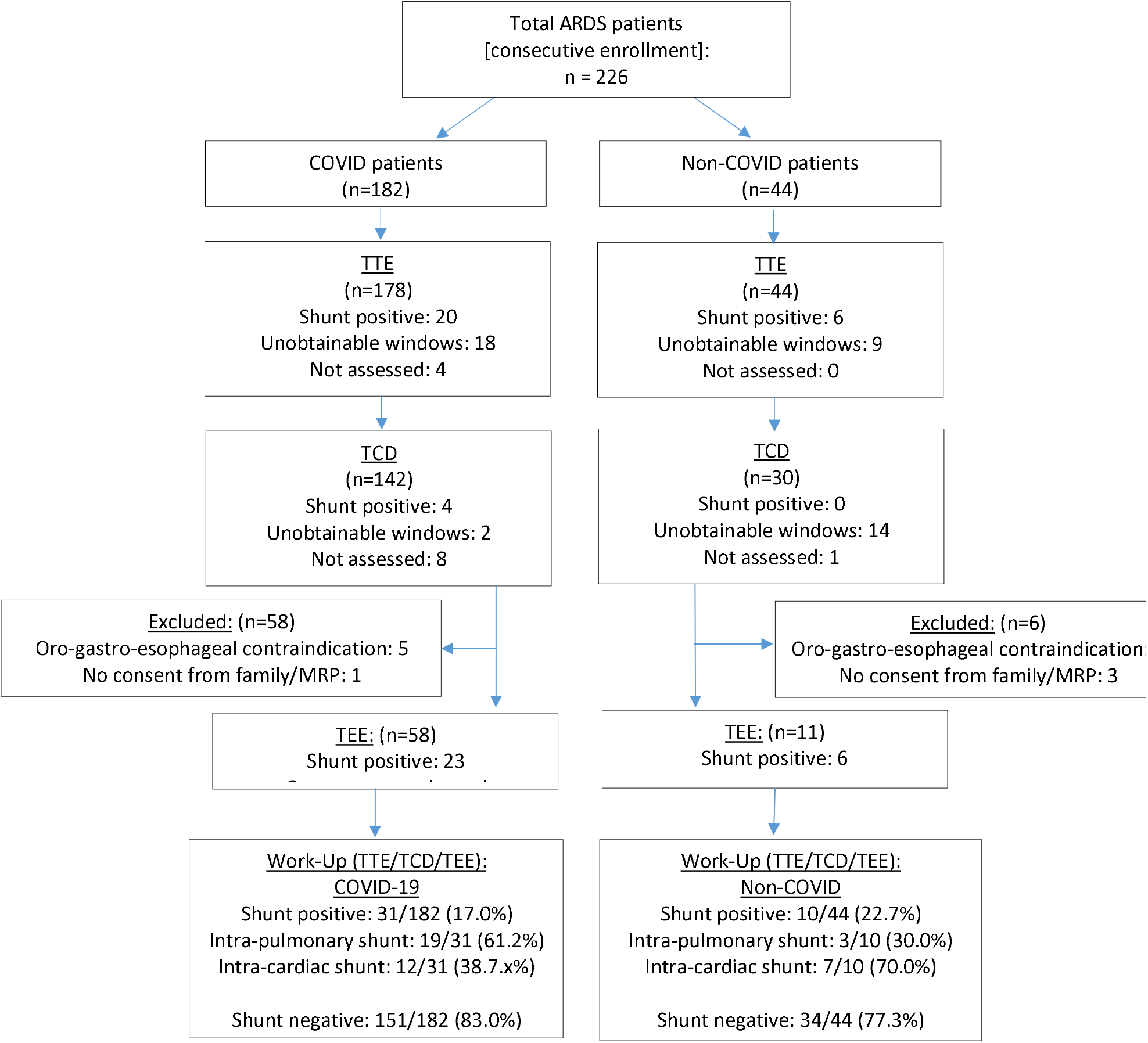
STROBE Diagram Flowchart for Hypoxemia Workup Assessing Shunt Presence. ARDS: acute respiratory distress syndrome; COVID-19: coronavirus disease-2019; STROBE (Strengthening the Reporting of Observational Studies in Epidemiology; MRP: most-responsible physician; TCD: transcranial Doppler; TEE: transesophageal echocardiography; TTE: transthoracic echocardiography

All patient investigations adhered to American Society of Echocardiography^11,14^ or American Society of Neuroimaging standards.^15,16^ All studies were supervised by board-certified echocardiographers or TCD sonographers from critical care/cardiac anesthesia/cardiology physicians. We performed external validation with over-readers of our TTE/TEE/TCD bubble studies, which allowed us to calculate inter-rater reliability (kappa statistic) to examine agreement in diagnosis of findings.

### Data collection

The Qpath database was queried for all TTE/TCD/TEE images/clips and reports. Demographic and clinical characteristic data were collected from registry databases within Alberta Health Services (eCritical/TRACER/DIMR) and included: COVID-19 status, age, sex, race/ethnicity, patient case-mix (medical, surgical, trauma), Acute Physiology And Chronic Health Evaluation (APACHE) II score,^17^ Charlson Comorbidity Index (CCI) score,^18^ respiratory mechanics (e.g. tidal volumes [TV], positive end-expiratory pressure [PEEP] static compliance, plateau pressures, PF ratio, arterial blood gas results (including A-a gradient), deadspace calculations (using arterial blood gas PCO2 compared to end-tidal CO2 from volumetric capnography), and type of ventilation at time of bubble study, types of interventions during hospital stay (e.g. prone positioning, airway-pressure release ventilation, pulmonary vasodilators, extracorporeal membrane oxygenation, renal replacement therapy), vasopressors/inotropes, steroid use, stress ulcer prophylaxis, venous thromboembolism (VTE) prophylaxis, ventilator-associated pneumonia (VAP) prophylaxis, sedation, analgesia, neuromuscular blockade use, and other baseline measures (e.g. vitals signs, lab values: complete blood count, troponin, D-dimer), where available.

The echocardiographic findings collected were: date of study, POCUS exam type (TTE or TEE) and location, presence/absence of intra-cardiac vs. intra-pulmonary shunt by bubble study, presence/absence of IAS defect by colour Doppler, and all other echocardiographic findings: e.g. biventricular size and function, valvulopathy, pericardial disease, superior or inferior vena caval size and respirophasic changes, etc. All study images were reviewed by at least 2 expert echocardiographers with National Board of Echocardiography certification to calculate inter-rater reliability for shunt identification. These POCUS assessors were blinded to clinical outcomes during the quality assurance oversight process. Treatment teams were not blinded to POCUS findings.

Clinical outcomes were reported through hospital discharge and at 90-days post-ICU admission. These included: ICU length of stay, hospital length of stay, duration of mechanical ventilation, and complications related to study procedures and hospitalization (Supplemental Appendix 2).

### Statistical analysis and sample size

Descriptive statistics were generated for baseline demographic, clinical characteristics, echocardiographic findings, and clinical outcome variables. Categorical data were summarized using frequency and column percentage and normal distributed data were described using mean and standard deviation. Non-normal distributed data were presented as median and inter-quartile ranges. Data were compared (where appropriate) using a Pearson’s chi-square test (categorical data), student t-test (normal distributed data), non-parametric Kruskal-Wallis test (non-normal distributed data). A *p* value <0.05 was considered statistically significant with 95% confidence intervals (CIs) also reported, if applicable. Missing data or lost-to-follow-up was <5%, so no imputation was required. There were no pre-specified sensitivity analyses or subgroups.

Inter-rater reliability for echocardiographic findings of shunt (intra-pulmonary vs. intra-cardiac) was calculated for Cohen’s kappa statistic, where the following interpretations were used: less than 0 (poor), 0-0.20 (slight), 0.21-0.40 (fair), 0.41-0.60 (moderate), 0.61-0.80 (substantial), 0.81-1.00 (almost perfect).^19,20^

Univariate logistic regression modelling was used to evaluate the association between unadjusted odds ratios (OR) with 95% CIs with mortality (continuous). Multivariable logistic regression modelling was also used to calculate adjusted ORs, adjusting for known variables including baseline demographics (age, sex) and clinical characteristics (Charlson comorbidity index) and (illness severity scores: e.g. APACHE II) to determine if the presence of shunt mortality exists after adjustment.

These statistical analyses were performed using Statistical Analysis System (SAS) Enterprise Guide 7.1 (Cary, NC, USA) or Microsoft Excel, version 14.0.6. All reporting of this observational cohort study was made in accordance with the STROBE (strengthening the reporting of observational studies in epidemiology) guidelines and checklist.^21^

In order to calculate study power, we used a reported incidence of shunts (e.g. PFOs) of approximately 19% in severe pneumonia/acute respiratory distress syndrome (ARDS), and a predicted increase in shunt of 15% with cor pulmonale (right-sided heart failure) physiology (up to a shunt rate of 34%).^7^ Using an alpha of 0.05 and power of 0.80, we calculated a minimum total sample size of 212 patients (106 patients per group). Considering an approximate attrition rate of 5%, this would require a minimum of 224 study participants for the incident shunt rate in the study.

## Results

### Demographics and clinical characteristics

We enrolled 226 patients. Of these, 182 were COVID-19 positive, and 44 composed the non-COVID control group (Figure 1). Baseline demographics and clinical characteristics are presented in Tables 1 and 2. Both groups had comparable tidal volumes, plateau pressures, and static compliance in keeping with high and equivalent rates of lung protective ventilation in both groups. The COVID-19 arm was associated with significantly higher rates of non-invasive positive pressure ventilation (NIPPV) and high flow nasal cannula oxygen administration (92.9% vs 77.3%, RD: 15.6%, 95% CI: 5.7-25.5, p=0.001) prior to intubation (Table 1). More patients in the COVID-19 arm underwent prone positioning (77.5% vs 43.2%, RD: 34.3%, 95% CI: 19.3-49.3, p=0.000004) (Table 3).

**Table 1.**
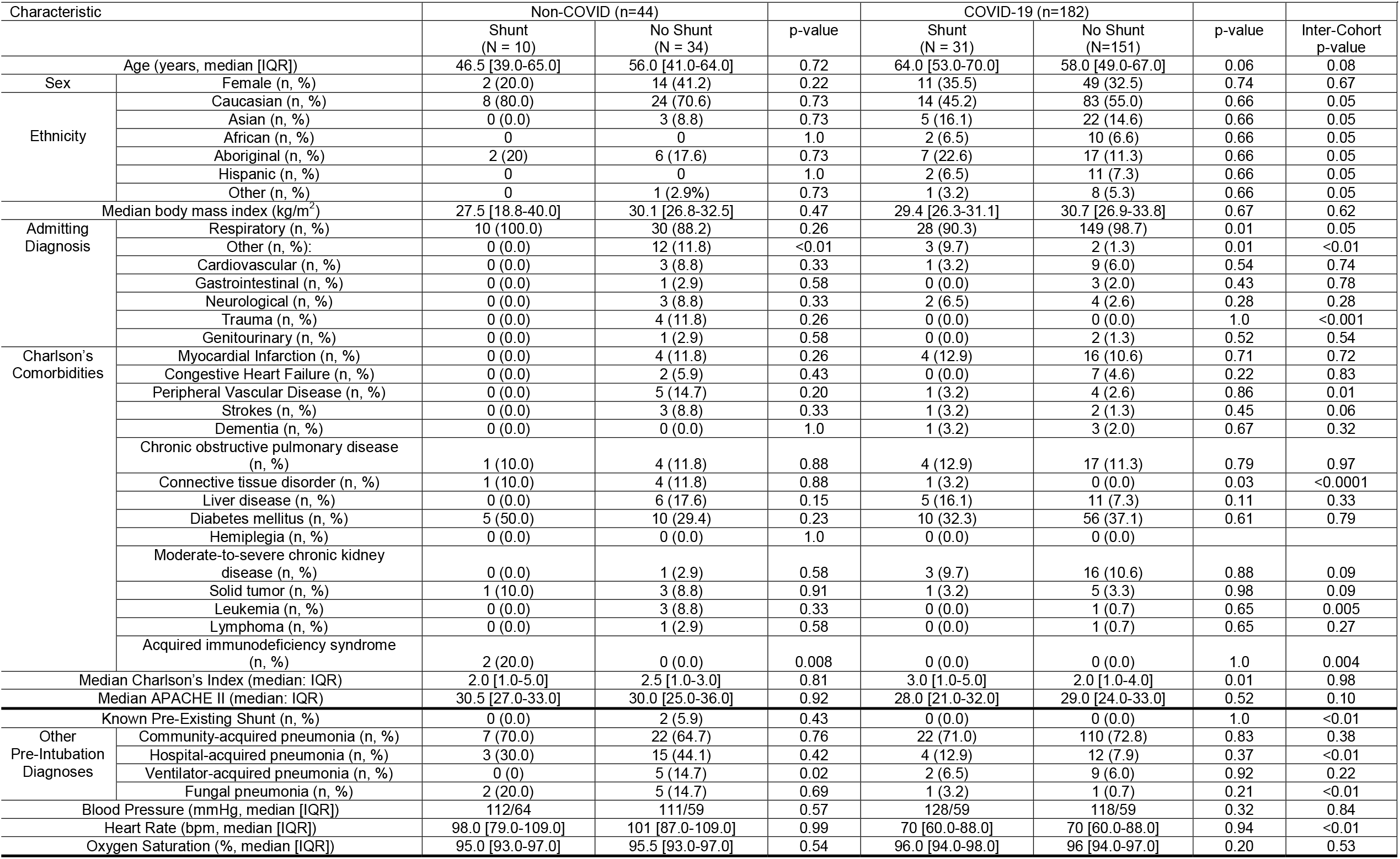
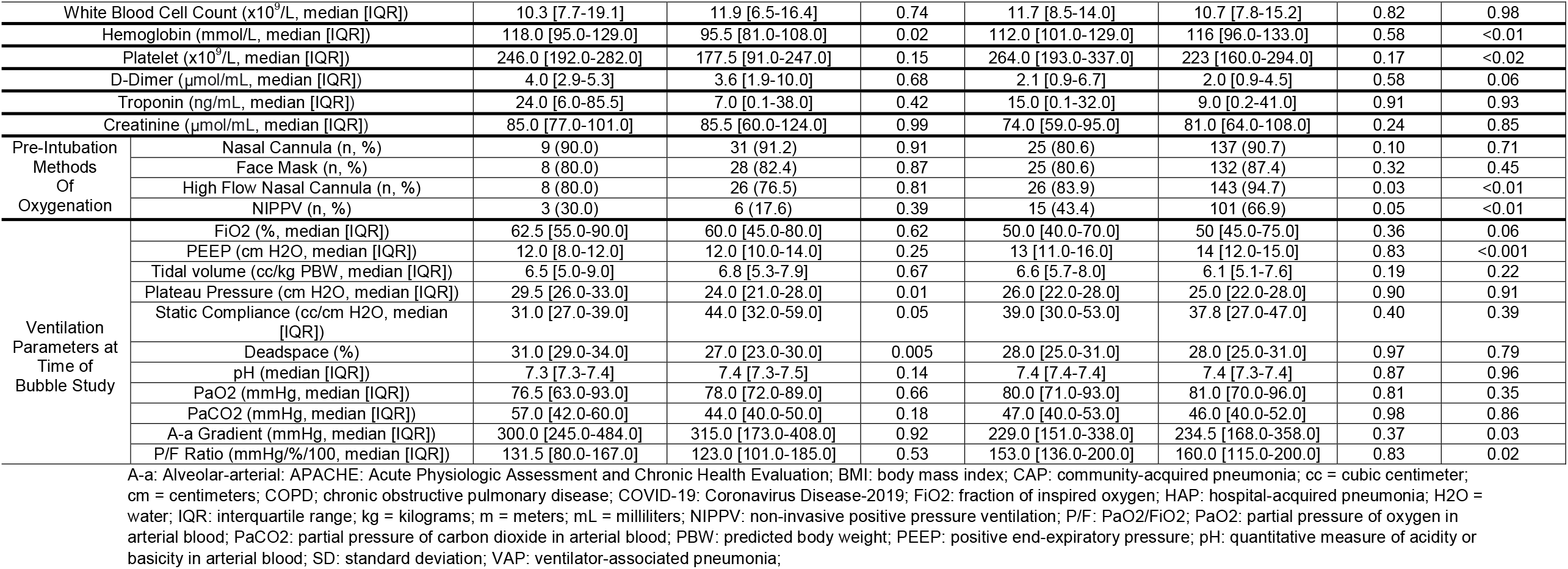
Baseline Demographics and Clinical Characteristics

**Table 2.** Ultrasonographic Findings (TTE/TCD ± TEE)

| Table 2. Ultrasonographic Findings (TTE/TCD ± TEE) |  |  |  |  |  |
| --- | --- | --- | --- | --- | --- |
| Finding |  | Non-COVID | COVID-19 | Risk Difference:<br>COVID vs. non-COVID (95% CI) | p-value |
| TTE Bubble Study | Positive (n, %) | 5/44 (11.4) | 20/178 (11.2) | -1.3 [-13.3 to 8.3] | 0.98 |
|  | Negative (with and without Valsalva) | 30/44 (68.2) | 136/178 (76.4) | +8.2 [-6.1 to 22.5] | 0.26 |
|  | Unable to obtain (n, %) | 9/44 (20.5) | 22/178 (12.4) | -8.1 [-19.6 to 3.4] | 0.17 |
| TCD Bubble Study | Positive (n, %) | 1/30 (3.3) | 4/142 (2.8) | -0.5 [-7.1 to 6.1] | 0.88 |
|  | Negative (with and without Valsalva) | 20/30 (66.7) | 128/142 (90.1) | +23.4 [9.7 to 37.1] | 0.0007 |
|  | Unable to obtain (n, %) | 9/30 (30.0) | 10/142 (7.0) | -23.0 [-10.7 to 35.3] | 0.0003 |
| TEE Bubble Study | Positive (n, %) | 6/11 (54.5) | 23/58 (39.7) | -14.8 [-46.6 to 17.0] | 0.36 |
|  | Negative (with and without Valsalva) | 5/11 (45.5) | 35/58 (60.3) | +14.8 [-17.0 to 46.6] | 0.36 |
| Total R-L Shunts (n, %) |  | 10/44 (22.7) | 31/182 (17.0) | -5.7 [-18.4 to 7.0] | 0.38 |
| Shunt Etiology | Intra-Cardiac (IAS defect, n, %) | 7/10 (70.0) | 12/31 (38.7) | -31.3 [-66.8 to 4.2] | 0.08 |
|  | PFO (n, %) | 7/10 (70.0) | 12/31 (38.7) | -31.3 [-66.8 to 4.2] | 0.08 |
|  | ASD (n, %) | 0/10 (0.0) | 0/31 (0.0) | 0.0 [0.0 to 0.0] | 1.0 |
|  | Intra-Pulmonary (n, %) | 3/10 (30.0) | 19/31 (61.2) | +31.2 [-4.4 to 66.8] | 0.08 |
| LV Function | Severely depressed (LVEF: <30%, n, %) | 4/44 (9.1) | 4/182 (2.2) | -6.9 [-13.0 to 8.1] | 0.73 |
|  | Depressed (LVEF: 30-50%, n, %) | 3/44 (6.8) | 17/182 (9.4) | +2.6 [-6.8 to 12.0] | 0.60 |
|  | Normal function (LVEF: 50-70%, n, %) | 35/44 (79.5) | 153/182 (89.0) | +9.5 [-1.5 to 20.5] | 0.47 |
|  | Hyperdynamic (LVEF: >70%, n, %) | 2/44 (4.5) | 8/182 (4.4) | -0.1 [-6.9 to 6.7] | 0.97 |
| RV Function | Severe dysfunction (n, %) | 2/44 (4.5) | 2/182 (1.0) | -3.5 [-7.7 to 1.0] | 0.12 |
|  | Moderate dysfunction (n, %) | 2/44 (4.5) | 6/182 (3.3) | -1.2 [-7.3 to 4.9] | 0.69 |
|  | Mild dysfunction (n, %) | 5/44 (11.4) | 18/182 (9.9) | -1.5 [-11.5 to 8.5] | 0.77 |
|  | Normal function (n, %) | 35/44 (79.5) | 155/182 (85.2) | +5.7 [-6.3 to 17.7] | 0.36 |
|  | Hyperdynamic (n, %) | 0/44 (0.0) | 1/182 (0.5) | 0.0 [0.0 to 0.0] | 1.0 |
| RV Size | Normal (n, %) | 36/44 (81.8) | 152/182 (83.5) | +1.7 [-10.6 to 14.0] | 0.79 |
|  | Mild dilation (RV < 2/3 LV size, n, %) | 1/44 (2.3) | 11/182 (6.0) | +3.7 [-3.7 to 11.1] | 0.32 |
|  | Moderate dilation (RV:LV size 1:1, n, %) | 6/44 (13.6) | 15/182 (8.2) | -5.4 [-14.9 to 4.1] | 0.27 |
|  | Severe dilation (RV > LV size, n, %) | 1/44 (2.3) | 4/182 (2.2) | -0.1 [-5.0 to 4.8] | 0.98 |
|  | Septal flattening (n, %) | 1/44 (2.3) | 2/182 (1.0) | -1.3 [-5.0 to 2.4] | 0.54 |
| Pericardial Effusion | No Effusion (n, %) | 36/44 (81.8) | 164/182 (90.1) | +8.3 [-2.2 to 18.8] | 0.12 |
|  | Trace/Small Effusion [0-1 cm] (n, %) | 7/44 (15.9) | 17/182 (9.3) | -6.6 [-16.7 to 3.5] | 0.20 |
|  | Moderate Effusion [1-2 cm] (n, %) | 1/44 (2.3) | 0/182 (0.0) | -2.3 [-4.5 to 0.1] | 0.19 |
|  | Large Effusion [>2 cm] (n, %) | 0/44 (0.0) | 1/182 (0.5) | +0.5 [-1.2 to 2.6] | 1.0 |
|  | Pericardial tamponade | 0/44 (0.0) | 1/182 (0.5) | +0.5 [-1.2 to 2.6] | 1.0 |
| Aortic valve | No aortic stenosis | 44/44 (100.0) | 179/182 (98.4) | -1.6 [-5.3 to 2.1] | 1.0 |
|  | Mild aortic stenosis | 0/44 (0.0) | 0/182 (0.0) | 0.0 [0.0 to 0.0] | 1.0 |
|  | Moderate aortic stenosis | 0/44 (0.0) | 2/182 (1.0) | +1.0 [-1.9 to 3.9] | 1.0 |
|  | Severe aortic stenosis | 0/44 (0.0) | 1/182 (0.5) | +0.5 [-1.2 to 2.6] | 1.0 |
|  | No aortic regurgitation | 42/44 (95.5) | 154/182 (83.5) | -12.0 [-0.5 to 23.5] | 0.06 |
|  | Trace-mild aortic regurgitation | 2/44 (4.5) | 26/182 (14.3) | +9.8 [-1.1 to 20.7] | 0.08 |
|  | Moderate aortic regurgitation | 0/44 (0.0) | 2/182 (1.0) | +1.0 [-1.9 to 3.9] | 1.0 |
|  | Severe aortic regurgitation | 0/44 (0.0) | 0/182 (0.0) | 0.0 [0.0 to 0.0] | 1.0 |
| Mitral valve | No mitral stenosis | 43/44 (97.7) | 179/182 (98.4) | +0.7 [-3.6 to 5.0] | 0.78 |
|  | Mild mitral stenosis | 1/44 (2.3) | 3/182 (1.6) | -0.7 [-5.0 to 3.6] | 0.78 |
|  | Moderate mitral stenosis | 0/44 (0.0) | 0/182 (0.0) | 0.0 [0.0 to 0.0] | 1.0 |
|  | Severe mitral stenosis | 0/44 (0.0) | 0/182 (0.0) | 0.0 [0.0 to 0.0] | 1.0 |
|  | No mitral regurgitation | 32/44 (72.7) | 137/182 (75.3) | +2.6 [-11.7 to 16.9] | 0.53 |
|  | Trace-mild mitral regurgitation | 12/44 (27.3) | 34/182 (18.7) | -8.6 [-21.9 to 4.7] | 0.20 |
|  | Moderate mitral regurgitation | 0/44 (0.0) | 10/182 (5.5) | +5.5 [-1.3 to 12.3] | 0.22 |
|  | Severe mitral regurgitation | 0/44 (0.0) | 1/182 (0.5) | +0.5 [-1.2 to 2.6] | 1.0 |
| Tricuspid valve | No tricuspid stenosis | 44/44 (100.0) | 182/182 (100.0) | 0.0 [0.0 to 0.0] | 1.0 |
|  | Mild tricuspid stenosis | 0/44 (0.0) | 0/182 (0.0) | 0.0 [0.0 to 0.0] | 1.0 |
|  | Moderate tricuspid stenosis | 0/44 (0.0) | 0/182 (0.0) | 0.0 [0.0 to 0.0] | 1.0 |
|  | Severe tricuspid stenosis | 0/44 (0.0) | 0/182 (0.0) | 0.0 [0.0 to 0.0] | 1.0 |
|  | No pulmonic regurgitation | 20/44 (45.5) | 85/182 (46.7) | +1.2 [-15.2 to 17.6] | 0.88 |
|  | Trace-mild tricuspid regurgitation | 19/44 (43.2) | 86/182 (47.2) | +4.0 [-12.4 to 20.4] | 0.63 |
|  | Moderate tricuspid regurgitation | 5/44 (11.4) | 11/182 (6.0) | -5.4 [-13.8 to 3.0] | 0.22 |
|  | Severe tricuspid regurgitation | 0/44 (0.0) | 0/182 (0.0) | 0.0 [0.0 to 0.0] | 1.0 |
| Pulmonic valve | No pulmonic stenosis | 44/44 (100.0) | 182/182 (100.0) | 0.0 [0.0 to 0.0] | 1.0 |
|  | Mild pulmonic stenosis | 0/44 (0.0) | 0/182 (0.0) | 0.0 [0.0 to 0.0] | 1.0 |
|  | Moderate pulmonic stenosis | 0/44 (0.0) | 0/182 (0.0) | 0.0 [0.0 to 0.0] | 1.0 |
|  | Severe pulmonic stenosis | 0/44 (0.0) | 0/182 (0.0) | 0.0 [0.0 to 0.0] | 1.0 |
|  | No pulmonic regurgitation | 42/44 (95.5) | 171/182 (94.0) | -1.5 [-9.1 to 6.1] | 0.70 |
|  | Trace-mild pulmonic regurgitation | 2/44 (4.5) | 11/182 (6.0) | +1.5 [-6.1 to 9.1] | 0.70 |
|  | Moderate pulmonic regurgitation | 0/44 (0.0) | 0/182 (0.0) | 0.0 [0.0 to 0.0] | 1.0 |
|  | Severe pulmonic regurgitation | 0/44 (0.0) | 0/182 (0.0) | 0.0 [0.0 to 0.0] | 1.0 |
| Pulmonary hypertension | No pulmonary hypertension | 35/44 (79.5) | 154/182 (84.6) | +5.1 [-7.1 to 17.3] | 0.41 |
|  | Mild pulmonary hypertension | 4/44 (9.1) | 15/182 (8.2) | -0.9 [-10.0 to 8.2] | 0.86 |
|  | Moderate pulmonary hypertension | 4/44 (9.1) | 12/182 (6.6) | -2.5 [-11.0 to 6.0] | 0.56 |
|  | Severe pulmonary hypertension | 1/44 (2.3) | 1/182 (0.5) | -1.8 [-4.8 to 1.2] | 0.27 |

ASD: atrial septal defect; CI: confidence interval; cm = centimeters; IAS: intra-atrial septal LV: left ventricle; LVEF: left ventricular ejection fraction; PFO: patent foramen ovale; RV: right ventricle; TEE: transesophageal echocardiography; TTE: transthoracic echocardiography;

**Table 3:**
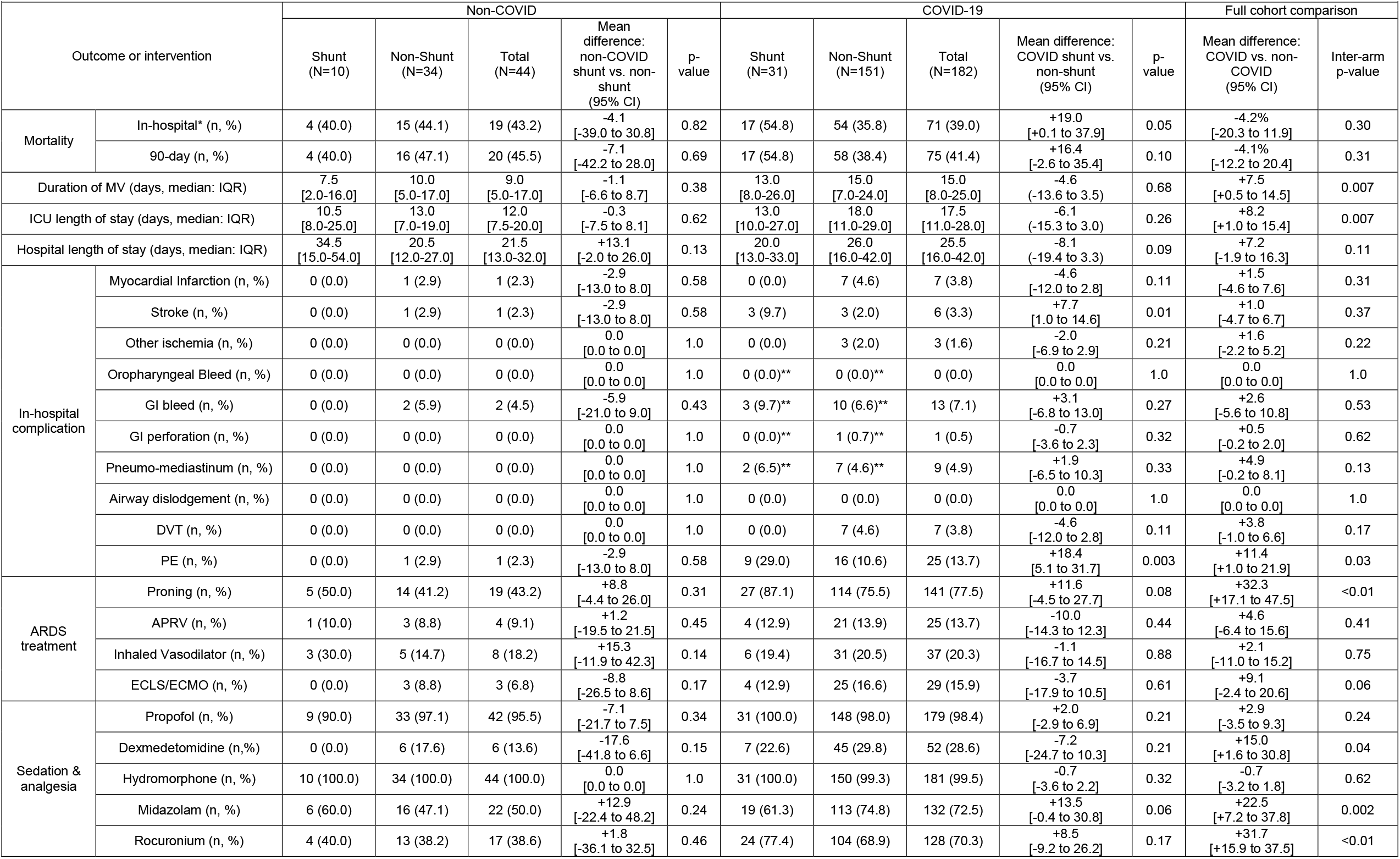
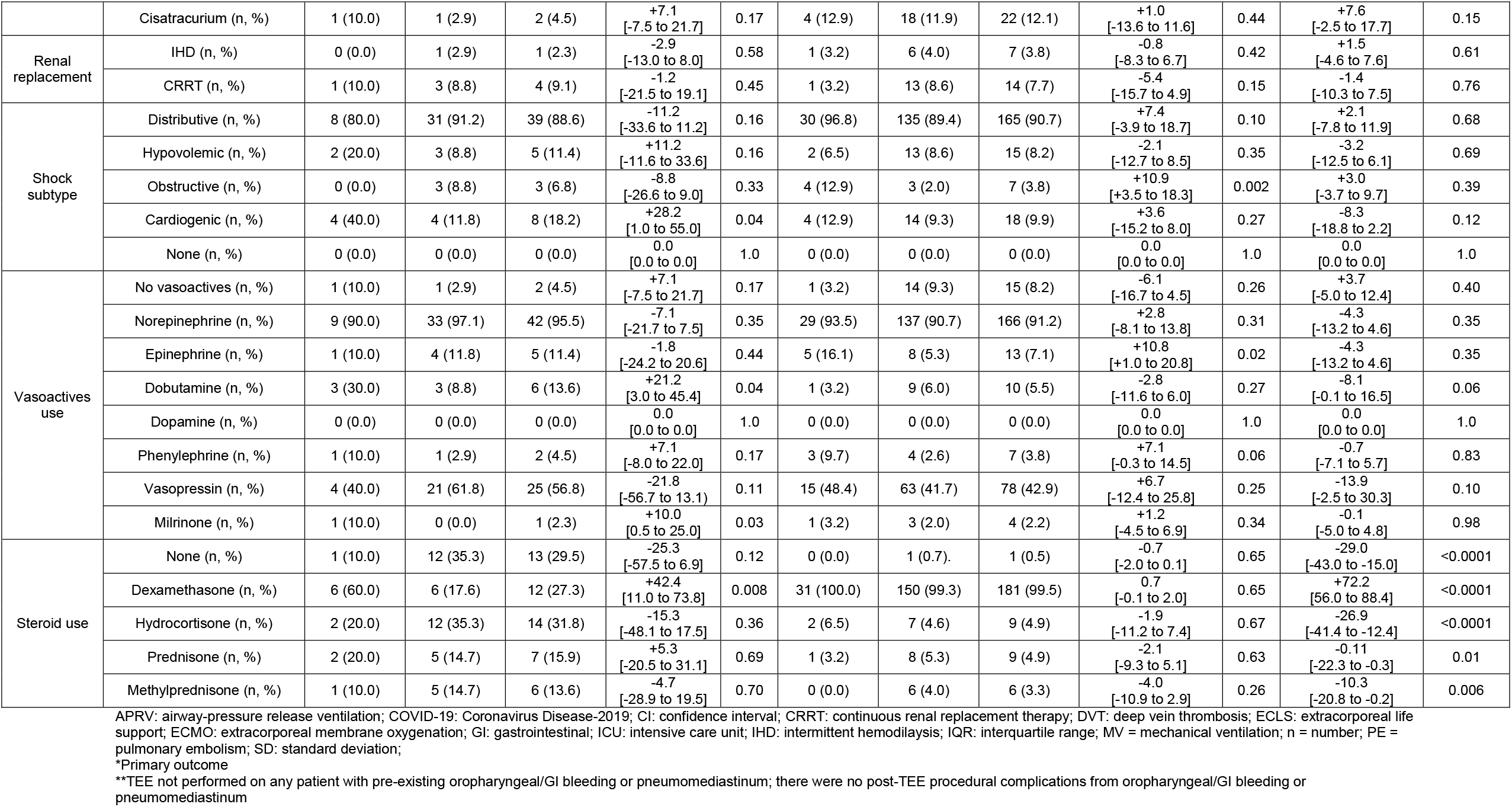
Outcomes and Co-Interventions

### Echocardiographic findings, right-to-left shunts and inter-rater reliability

Echocardiographic findings and the percentage with a right-to-left shunt are shown in Table 3. In the COVID-19 group, 31/182 patients (17.0%) had a shunt identified, of which 12 were intra-cardiac (38.7%) and 19 (61.3%) were intra-pulmonary shunts. In the non-COVID group, 10/44 (22.7%) of patients had an identified shunt; of which 7 (70.0%) were intra-cardiac shunts and 3 (30.0%) were intra-pulmonary. There was no statistically significant difference in the overall rate of shunt between the COVID-19 and non-COVID groups (17.0% vs 22.7%, risk difference [RD]: -5.7%, 95% CI: -18.4-7.0, p=0.38). There was a non-significant higher proportion of intra-pulmonary shunts in the COVID-19 group compared to non-COVID (61.2% vs 30.0% respectively, RD: 31.2%, 95% CI: -4.4-66.8%, p=0.08) (Table 2). Inter-rater reliability was high for TTE/TCD/TEE shown in Supplemental Table 1.

### Clinical Outcomes

Clinical outcomes are summarized in Table 3. For the primary outcome, there was higher in-hospital mortality among COVID shunt patients compared to no shunt (54.8% vs 35.8%, RD: 19.0%, 95% CI: 0.1-37.9, p=0.05). However, this difference was no longer significant at 90-day mortality (54.8% vs 38.4%, RD: 16.4%, 95% CI: -2.6-35.4%, p=0.10). There was no difference in either in-hospital (39.0% vs 43.2%, RD: -4.2%, 95% CI: -20.3-11.9%, p=0.30) or 90-day mortality (41.2% vs 45.5%, RD: -4.1%, 95% CI: -12.2-20.4%, p=0.31) between the COVID-19 and non-COVID arms.

COVID-19 infection was associated with a significantly longer median duration of mechanical ventilation (15.0 days [IQR: 8.0-25.0] vs 9.0 days [IQR: 5.0-17.0], RD: 7.5 days, 95% CI: 0.5-14.5, p=0.007) as compared to the control group. There was also a longer median ICU length of stay (17.5 days [IQR: 11.0-28.0] vs 12.0 days [IQR: 7.5-20.0], RD: 8.2 days, 95% CI: 1.0-15.4, p=0.007).

There was no measurable increase in complications attributable to performing TEE (oropharyngeal/GI bleeding or pneumomediastinum). There were no detected cases of esophageal perforation (Table 3).

### Kaplan-Meier Curves and multivariable logistic regression analysis

Kaplan-Meier curves for 90-day mortality are presented in Figure 2. There was a significant difference in shunt vs. no shunt in COVID patients (p=0.04) [Figure 2c]. The remaining log-rank tests were not significant and after adjustment for multiple comparisons.

**Figure 2.**
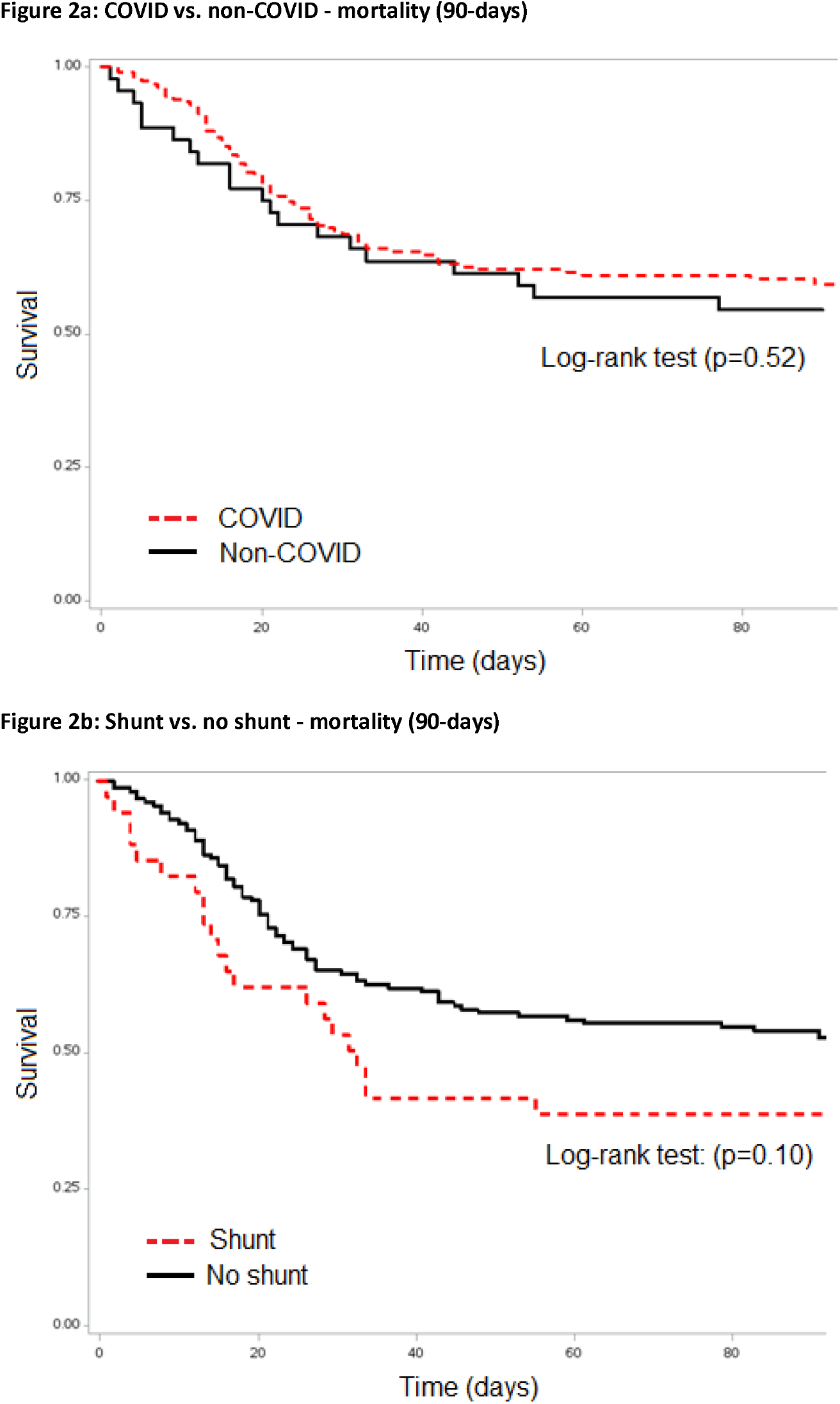
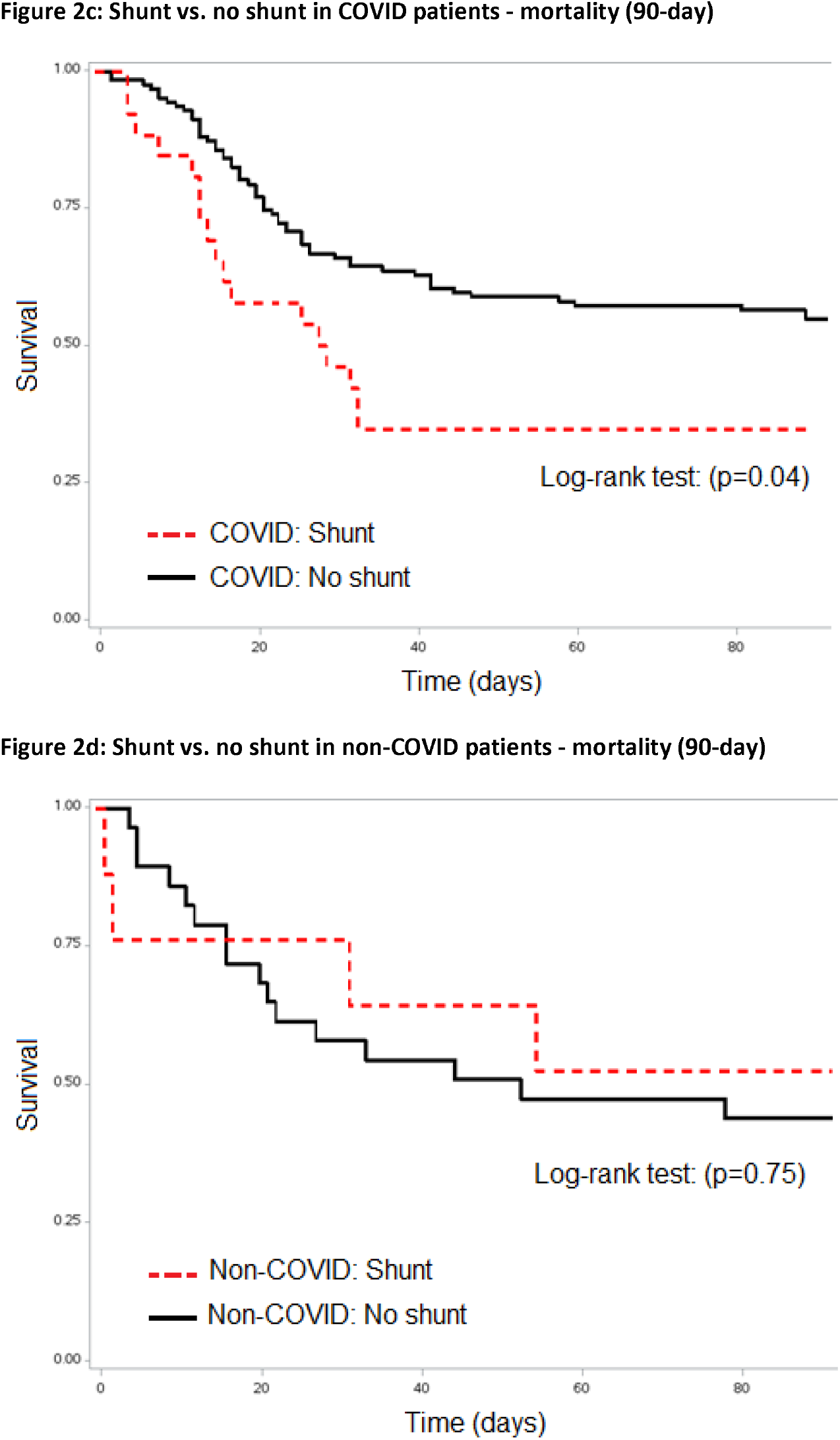
Kapan-Meier Curves

The combined regression analysis in the full cohort showed no significant difference in 90-day mortality based on the presence of any shunt or in the intra-cardiac and intra-pulmonary shunt subtypes. The regression adjusting for Charlson’s Health Score specifically did show a significant increase in 90-day mortality in the shunt portion of the overall cohort (OR: 1.28, 95% CI: 1.09-1.52). No other individual co-variable adjustments showed a significant signal for increased mortality in any group (Table 4).

## Discussion

In this study, COVID-19 shunt rates were not significantly different compared to non-COVID ARDS. Our findings align with the recent meta-analysis,^12^ suggesting approximately ∼1 in 5 patients with ARDS had a R-L shunt. We also found an association between R-L shunts and increased hospital mortality, but this was no longer significant at 90-day mortality, or after multivariable adjustment. An updated Forest plot for pooled analysis with our systematic review may suggest a statistically significant increase in mortality (OR: 1.26, 95% CI: 1.06-1.49, p=0.009) (Supplemental Figure 3).

Given that approximately 1-in-5 patients with ARDS may have a R-L shunt, this study is a reminder to clinicians to consider screening patients, and if present, then to consider targeted therapies. This study also highlights that not all R-L shunts are intra-pulmonary, and that different shunts will have different treatment implications. Specifically, intra-pulmonary shunts are most often due to abnormal vasodilation of pulmonary vessels. Therefore, treatment focuses on: (1) reducing underlying inflammation/infection leading to pulmonary vasodilation (e.g. corticosteroids);^22^ (2) vigilant PEEP titration and ventilator optimization, to prevent over dilation of pulmonary vessels, while preventing shunt from atelectasis from occurring;^23,24^ and, (3) avoiding pulmonary vasodilators (e.g. epoprostenol, nitric oxide, sildenafil).^25,26^ In contrast, intra-cardiac shunt management should lower right-sided heart pressures to prevent further shunting through an intra-atrial septum defect (e.g. PFO or ASD). Treatments include: (1) pulmonary vasodilators (e.g. inhaled nitric oxide, epoprostenol) and/or inodilators (e.g. milrinone, dobutamine) through reducing RV afterload and improving RV function;^23,27,28^ (2) lowering ventilator settings (e.g. PEEP, plateau pressures);^23,27,28^ (3) closure or repair of an intra-septal defects (PFO, ASD) to prevent further R-L shunting;^23,27,28^; and (4) diuresis to offload RV volume overload.^23,27,28^ Regardless, diagnosing shunt in ARDS patients starts with high suspicion and prompt diagnosis.

Guidelines have promoted standardizing ARDS management, like using low-tidal volume ventilation^29^ and proning,^30^. among other strategies. There has also been adoption of higher PEEP in both COVID and non COVID-ARDS. Our study is a reminder that indiscriminate use of PEEP or pulmonary vasodilators may be harmful in the wrong patient. Future work could include: (1) identifying which patients to screen for shunt; and, (2) potential interventions to reduce mortality from shunts.

This study has its strengths. We confirmed that research is still feasible in the midst of a pandemic, by undertaking the largest study of shunts in COVID-19 ARDS. We have designed an extensive protocol for ICU shunt work-up, which was performed by intensivists during the pandemic, saving on personal protective equipment. We investigated different shunt types, co-interventions, and duration of mechanical ventilation plus other respiratory adjuncts, which is not routinely reported in ARDS literature.^12^ Our study reinforces the safety of intensivist and trainee TEE, given that there were no procedural complications,^31^ and our inter-relator scores highlight that ICU echocardiography and TCD is feasible and reliable.^32,33^ We performed both unadjusted and adjusted ORs analysis using multivariable logistic regression to account for known confounders (e.g age, illness severity), in keeping with STROBE and Newcastle-Ottawa score recommendations.^21,34^

There are several limitations to this study. The smaller size of our non-COVID arm, leading to imbalance and potential loss of statistical power; however, given the higher number of COVID-19 vs. non-COVID patients, this shunt data is representative of the ICU population at the time. Patient factors such as obesity and poor windows affected our ability to perform shunt fractions, even with TEE. Finally, the relatively low rate of right ventricular dysfunction is intriguing, and may be because these patients underwent ultrasonographic assessments early in their course on the ventilator.

### Conclusion

There was no evidence of increased R-L shunt rates in COVID-19 compared to non-COVID and historical controls. Right-to-left shunt presence was associated with increased in-hospital mortality for COVID-19 patients, but this did not persist for 90-day mortality or after adjusting using logistic regression.

## Data Availability

All data produced in the present study are available upon reasonable request to the authors

## Acknowledgements

We are grateful for the assistance and support from the University of Alberta Department of Critical Care Medicine Research Office, the University of Alberta and Royal Alexandra Hospital, Mazankowski Heart Institute and Grey Nuns Hospital Intensive Care Units staff, nurses and respiratory therapists, and all the University of Alberta Critical Care Ultrasound Service rotators.

## Author Contributions

Vincent Lau, Graham Mah, Andrea Robinson, Leon Byker, Lazar Milovanovic, Aws Alherbish, Jeffrey Odenbach, Cristian Vadeanu, David Lu, Leo Smyth, Mitchell Rohatensky, Dustin Anderson, Sanam Verma, Jocelyn Slemko, Peter Brindley, Jim Kustogiannis, Michael Jacka, Andrew Shaw, Jonathan Windram, Matt Wheatley, Xiaoming Wang, Oleksa Rewa, Sean M. Bagshaw, and Brian Buchanan have: (1) made substantial contributions to conception and design, acquisition of data, analysis and interpretation of data; (2) drafted the submitted article and revised it critically for important intellectual content, and (3) provided final approval of the version to be published.

### Conception

Lau, Brindley, Kutsogiannis, Jacka, Rewa, Bagshaw, Buchanan

### Background

Lau, Mah, Lu, Alherbish, Brindley, Kutsogiannis, Jacka, Odenbach, Anderson Shaw, Windram, Wheatley, Rewa, Bagshaw, Buchanan

### Design

Lau, Alherbish, Brindley, Kutsogiannis, Jacka, Anderson, Rewa, Bagshaw, Buchanan

Acquisition of data: Lau, Mah, Robinson, Byker, Milovanovic, Alherbish, Odenbach, Vadeanu, Lu, Smyth, Rohatensky, Anderson, Verma, Slemko

### Analysis of data

Lau, Mah, Robinson, Byker, Milovanovic, Alherbish, Odenbach, Vadeanu, Lu, Smyth, Rohatensky, Anderson, Verma, Slemko, Brindley, Kustogiannis, Jacka, Shaw, Windram, Wheatley, Wang, Rewa, Bagshaw, Buchanan

### Drafting the manuscript

Lau, Mah, Robinson, Byker, Milovanovic, Alherbish, Odenbach, Vadeanu, Lu, Smyth, Rohatensky, Anderson, Verma, Slemko, Brindley, Kustogiannis, Jacka, Shaw, Windram, Wheatley, Wang, Rewa, Bagshaw, Buchanan

### Revising the manuscript

Lau, Mah, Robinson, Byker, Milovanovic, Alherbish, Odenbach, Vadeanu, Lu, Smyth, Rohatensky, Anderson, Verma, Slemko, Brindley, Kustogiannis, Jacka, Shaw, Windram, Wheatley, Wang, Rewa, Bagshaw, Buchanan

## Supplements

Supplemental Appendix 1: STROBE Checklist

Supplemental Appendix 2: Protocol and Operational Definitions

Supplemental Figure 1: COVID Shunt Protocol Bubble Studies

Supplemental Table 1: Kappa Inter-Rater Reliability

Supplemental Figure 2: Right-to-Left Shunt Percent Positivity Rate

Supplemental Table 2: Multivariable Logistic Regression

Supplemental Figure 3: Updated Forest Plot (meta-analysis)

