## Supplemental Appendix 2 for "Intra-pulmonary and intra-cardiac shunts in adult COVID-19 versus non-COVID ARDS ICU patients using echocardiography and contrast bubble studies (COVID-Shunt Study): a prospective, observational cohort study"

During TTE, ECG leads were placed where possible, and we used the cardiac preset with the phased-array probe. Full echocardiography was otherwise performed, where available. An apical 4-chamber or subcostal long axis view (SubX LAX) was obtained to screen for intra-cardiac/intra-pulmonary shunt. Colour Doppler was placed over the intra-atrial septum at varying Nyquist levels to screen for an intra-atrial septum (IAS) defect from a SubX LAX view. Agitated-saline bubble studies were injected using a 3-way Luer-lock system and 10 mL saline syringes. Via either central line or peripheral intravenous we mixed and injected 0.5mL of patient’s blood, 0.5-1mL of air, and 10 mL of saline. We recorded for 30 second (representing >10 cardiac cycles) during each bubble study. We recorded at least 1-2 beats of pre-injection cardiac cycles prior to bubble injection for the purpose of counting the cardiac cycle. Bubble studies were performed at least twice (without and with simulated Valsalva).^1,2^

        	If the TTE bubble study was negative, we followed with a TCD bubble study, given its increased sensitivity in detecting R-L shunts. For TCD, we used the ultrasound machine with a phased-array probe in transcranial preset. The transtemporal window was insonated with a pulse-wave spectral Doppler on the ipsilateral middle cerebral artery (MCA) at the M1 segment (3.5-5.5 mm gate) or another intra-cranial artery, if the MCA was not detectable. The same agitated saline protocol was used, as described above.^3,4^

        	TEE was performed if either the TTE or TCD bubble studies were positive, or if either study was indeterminate or unattainable (technically difficult study). Full standard TEE views were recorded. Both 2D and Colour Doppler were used to interrogate the IAS for intra-cardiac shunt defects. The main views were the mid-esophageal 4-chamber, and mid-esophageal bicaval view. The agitated saline protocol was the same as described above. Colour Nyquist was adjusted from the highest to the lowest possible levels to detect the presence of intra-cardiac shunt in the IAS, either patent foramen ovale (PFO) or atrial septal defects (ASD).^1,2^

Four Canadian intensive care units participated in the study: the University of Alberta Hospital General Systems ICU, the Mazankowski Cardiovascular ICU, the Royal Alexandra Hospital ICU, and the Grey Nuns Hospital ICU.

Operational Definitions:

Myocardial infarction was defined as acute myocardial injury with clinical evidence of acute myocardial ischemia and with detection of a rise and/or fall of cardiac troponin values with at least 1 value about the 99^th^ percentile upper limit of lab testing and at least 1 of the following: symptoms of myocardial ischemia; new ischemic ECG changes; development of pathological Q waves; imaging evidence of new loss of viable myocardium or new regional wall motion abnormality in a pattern consistent with an ischemic etiology; identification of a coronary thrombus by angiography or autopsy [Thygesen Circulation 2018].^5^

Cardio-respiratory arrest was defined as the cessation of effective ventilation and circulation.^6^

Stroke was defined as central nervous system infarction of the brain, spinal cord, or retinal cell death attributable to ischemia, based on: pathological imaging; or other objective evidence of cerebral, spinal cord or retinal focal ischemic injury in a defined vascular distribution; or, clinical evidence of cerebral, spinal cord, or retinal focal ischemic injury based on symptoms on symptoms persisting ≥24 hours or until death, and other etiologies excluded.^7^

Systemic ischemic event was defined as a thrombosis or embolism which originate or travel through the systemic arterial circulation.^8^

Oropharyngeal/gastrointestinal bleeding/perforation was defined as either dental trauma, submucosal hematoma of pharyngeal area, jaw subluxation, oropharyngeal bleeding, gastro-esophageal perforation following TEE probe insertion.^9^

Pneumomediastinum was defined as the presence of air in the mediastinum. This condition can result from physical trauma or other situations that lead to air escaping from the lungs, airways or bowel into the chest cavity. Pneumomediastinum is a rare situation and occurs when air leaks into the mediastinum.^10^

Respiratory endotracheal tube dislodgement was defined as an ETT moving at least 2 cm, or unplanned extubation.^11^

Venous thromboembolism (VTE) was defined as: blood clots in veins, which can manifest as deep vein thrombosis (in the lower extremities) and pulmonary embolism (in the lungs).^12^
