## Supplemental Figure 1 for "Intra-pulmonary and intra-cardiac shunts in adult COVID-19 versus non-COVID ARDS ICU patients using echocardiography and contrast bubble studies (COVID-Shunt Study): a prospective, observational cohort study"

Supplemental Figure 1. COVID-19 Pneumonia Hypoxemia Shunt Protocol

All adult ICU patients

(intubated/mechanical ventilation)

Screen for R-L shunt with contrast-enhanced TTE

(bubble study with and without Valsalva)

*Negative (-)
bubble study*

**Positive (+)
bubble study
(or no windows)**

**Positive (+)
bubble study**

**or no windows**

Assess for IAS defect (PFO/ASD) with contrast-enhanced TEE

(bubble study with and without Valsalva)

Assess for R-L shunt with contrast-enhanced TCD (bubble study)

**Bubbles cross within 4-8 cardiac cycles & no evidence of IAS defect**

**Bubbles cross within 1-2 cardiac cycles & evidence of IAS defect**

*Negative (-)
bubble study*

No evidence of shunt

Intra-cardiac shunt

Intra-pulmonary shunt

ASD: atrial septal defect, COVID-19: coronavirus disease 2019, IAS: intra-atrial septal defect, ICU: intensive care unit, R-L: shunt, TCD: transcranial Doppler; TEE: trans-esophageal echocardiography, TTE: trans-thoracic echocardiography
