## Supplemental Figure 3 for "Intra-pulmonary and intra-cardiac shunts in adult COVID-19 versus non-COVID ARDS ICU patients using echocardiography and contrast bubble studies (COVID-Shunt Study): a prospective, observational cohort study"

Supplemental Figure 3. Mortality Forest Plot of R-L shunt vs. no shunt ARDS patients

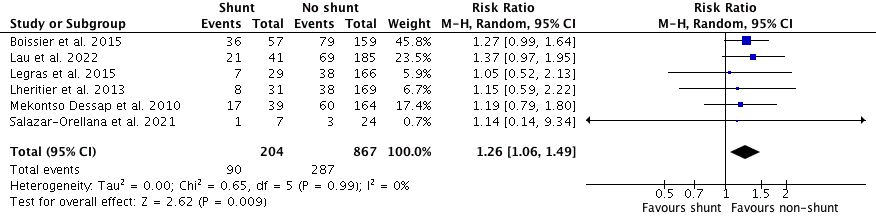

ARDS: acute respiratory distress syndrome; COVID-19: coronavirus disease-2019; R-L: right-to-left-shunt
