## Supplemental Table 1 for "Intra-pulmonary and intra-cardiac shunts in adult COVID-19 versus non-COVID ARDS ICU patients using echocardiography and contrast bubble studies (COVID-Shunt Study): a prospective, observational cohort study"

**Supplemental Table 1:** Inter-rater reliability (kappa) of right-to-left shunt and intra-pulmonary vs. intra-cardiac shunts using TTE/TCD/TEE and contrast bubble studies (all studies)

| Positive bubble study (TTE/TEE) |  | | |
| --- | --- | --- | --- |
| Inter-relater reliability | Rater #2 | | |
| Rater #1 | Positive bubble study | Negative bubble study | Total |
| Positive bubble study | 35 | 2 | 37 |
| Negative bubble study | 0 | 189 | 189 |
| Total | 35 | 191 | 226 |
| Rater agreement (observed) | 35 | 189 | 224 |
| Rater agreement by chance (expected) | 5.73 | 159.06 | 163.79 |
|  | Kappa | 95% CI (lower limit) | 95% CI (upper limit) |
| Cohen’s kappa statistic | 0.968 | 0.923 | 1.00 |
| Positive bubble study (TCD) |  |  |  |
| Inter-relater reliability | Rater #2 | | |
| Rater #1 | Positive bubble study | Negative bubble study | Total |
| Positive bubble study | 2 | 0 | 2 |
| Negative bubble study | 0 | 189 | 189 |
| Total | 2 | 189 | 191 |
| Rater agreement (observed) | 2 | 189 | 191 |
| Rater agreement by chance (expected) | 0.021 | 187.02 | 187.04 |
|  | Kappa | 95% CI (lower limit) | 95% CI (upper limit) |
| Cohen’s kappa statistic | 1.00 | 1.00 | 1.00 |
| PFO or ASD identified (TTE/TEE) | Rater #2 |  |  |
| Rater #1 | PFO/ASD present | PFO/ASD absent | Total |
| PFO/ASD present | 21 | 1 | 22 |
| PFO/ASD absent | 5 | 193 | 198 |
| Total | 26 | 194 | 220 |
| Rater agreement (observed) | 21 | 193 | 214 |
| Rater agreement by chance (expected) | 2.10 | 173.1 | 175.8 |
|  | Kappa | 95% CI (lower limit) | 95% CI (upper limit) |
| Cohen’s kappa statistic | 0.864 | 0.749 | 0.970 |

ASD: atrial septal defect; CI = confidence interval; PFO: patent foramen ovale; TCD: transcranial Doppler; TEE: transesophageal echocardiography; TTE: transthoracic echocardiography;
