## Supplemental Table 2 for "Intra-pulmonary and intra-cardiac shunts in adult COVID-19 versus non-COVID ARDS ICU patients using echocardiography and contrast bubble studies (COVID-Shunt Study): a prospective, observational cohort study"

**Table 4:** Multivariable logistic regression (unadjusted and adjusted odds ratios)

|  | **90-Day Mortality** | | |
| --- | --- | --- | --- |
| **Risk Factor** | **Prevalence (n=226, %)** | **Unadjusted Odds Ratio (95% CI)** | **Adjusted Odds Ratio (95% CI)** |
| Intubated ARDS (any shunt) | 41 (18.1) | 1.56 (0.79-3.08) | 1.22 (0.57-2.57)** |
| - Adjusted for age |  |  | 1.01 (0.99-1.04) |
| - Adjusted for sex |  |  | 1.17 (0.62-2.18) |
| - Adjusted for APACHE |  |  | 1.02 (0.98-1.06) |
| - Adjusted for Charlson’s |  |  | 1.28 (1.09-1.52) |
| Intubated ARDS (intra-cardiac shunt) | 19 (8.4) | 1.58 (0.62-4.06) | 1.78 (0.64-4.94)** |
| - Adjusted for age |  |  | 1.02 (0.99-1.04) |
| - Adjusted for sex |  |  | 1.18 (0.63-2.22) |
| - Adjusted for APACHE |  |  | 1.01 (0.98-1.05) |
| - Adjusted for Charlson’s |  |  | 1.29 (1.09-1.52) |
| Intubated ARDS (intra-pulmonary shunt) | 22 (9.7) | 1.42 (0.59-3.42) | 0.82 (0.30-2.24)** |
| - Adjusted for age |  |  | 1.02 (0.99-1.04) |
| - Adjusted for sex |  |  | 1.17 (0.62-2.18) |
| - Adjusted for APACHE |  |  | 1.01 (0.98-1.06) |
| - Adjusted for Charlson’s |  |  | 1.29 (1.09-1.52) |
| **Risk Factor** | **Prevalence (n=182, %)** | **Unadjusted Odds Ratio (95% CI)** | **Adjusted Odds Ratio (95% CI)** |
| COVID-19 ARDS (any shunt) | 31 (17.0) | 1.96 (0.88-4.20) | 1.53 (0.64-3.67)** |
| - Adjusted for age |  |  | 1.02 (0.99-1.06) |
| - Adjusted for sex |  |  | 0.77 (0.37-1.60) |
| - Adjusted for APACHE |  |  | 1.06 (1.01-1.11) |
| - Adjusted for Charlson’s |  |  | 1.15 (0.95-1.40) |
| COVID-19 ARDS (intra-cardiac shunt) | 12 (6.6) | 3.04 (0.88-10.51) | 2.83 (0.75-10.70)** |
| - Adjusted for age |  |  | 1.03 (0.99-1.06) |
| - Adjusted for sex |  |  | 0.81 (0.39-1.67) |
| - Adjusted for APACHE |  |  | 1.05 (1.00-1.10) |
| - Adjusted for Charlson’s |  |  | 1.16 (0.96-1.41) |
| COVID-19 ARDS (intra-pulmonary shunt) | 19 (10.4) | 1.31 (0.59-3.42) | 0.93 (0.31-2.80)** |
| - Adjusted for age |  |  | 1.03 (0.99-1.06) |
| - Adjusted for sex |  |  | 0.79 (0.38-1.63) |
| - Adjusted for APACHE |  |  | 1.05 (1.01-1.10) |
| - Adjusted for Charlson’s |  |  | 1.16 (0.95-1.40) |
| **Risk Factor** | **Prevalence (n=44, %)** | **Unadjusted Odds Ratio (95% CI)** | **Adjusted Odds Ratio (95% CI)** |
| Non-COVID ARDS (any shunt) | 10 (22.7) | 0.75 (0.18-3.14) | 0.75 (0.08-6.79)** |
| - Adjusted for age |  |  | 0.98 (0.89-1.08) |
| - Adjusted for sex |  |  | 3.65 (0.67-20.04) |
| - Adjusted for APACHE |  |  | 0.85 (0.74-0.97) |
| - Adjusted for Charlson’s |  |  | 2.64 (1.12-6.62) |
| Non-COVID ARDS (intra-cardiac shunt) | 7 (15.9) | 0.42 (0.07-2.46) | 0.37 (0.03, 5.37)** |
| - Adjusted for age |  |  | 0.98 (0.89-1.07) |
| - Adjusted for sex |  |  | 3.87 (0.70-21.47) |
| - Adjusted for APACHE |  |  | 0.85 (0.74-0.97) |
| - Adjusted for Charlson’s |  |  | 2.63 (1.17-5.91) |
| Non-COVID ARDS (intra-pulmonary shunt) | 3 (6.8) | 2.56 (0.21-30.57) | 9.96 (0.07-1419.34)** |
| - Adjusted for age |  |  | 0.95 (0.85-1.07) |
| - Adjusted for sex |  |  | 5.04 (0.79-32.05) |
| - Adjusted for APACHE |  |  | 0.85 (0.75-0.97) |
| - Adjusted for Charlson’s |  |  | 3.08 (1.19-7.98) |

**adjusted for: patient’s age, sex, APACHE II, Charlson’s Cormorbidity Index (combined)

APACHE: Acute Physiologic Assessment and Chronic Health Evaluation; ARDS: acute respiratory distress syndrome; CI = confidence interval; COVID-2019: Coronavirus Disease-2019; n = number
